# Projected resurgence of COVID-19 in the United States in July—December 2021 resulting from the increased transmissibility of the Delta variant and faltering vaccination

**DOI:** 10.1101/2021.08.28.21262748

**Authors:** Shaun Truelove, Claire P. Smith, Michelle Qin, Luke C. Mullany, Rebecca K. Borchering, Justin Lessler, Katriona Shea, Emily Howerton, Lucie Contamin, John Levander, Jessica Salerno, Harry Hochheiser, Matt Kinsey, Kate Tallaksen, Shelby Wilson, Lauren Shin, Kaitlin Rainwater-Lovett, Joseph C. Lemaitre, Juan Dent, Joshua Kaminsky, Elizabeth C. Lee, Javier Perez-Saez, Alison Hill, Dean Karlen, Matteo Chinazzi, Jessica T. Davis, Kunpeng Mu, Xinyue Xiong, Ana Pastore y Piontti, Alessandro Vespignani, Ajitesh Srivastava, Przemyslaw Porebski, Srinivasan Venkatramanan, Aniruddha Adiga, Bryan Lewis, Brian Klahn, Joseph Outten, James Schlitt, Patrick Corbett, Pyrros Alexander Telionis, Lijing Wang, Akhil Sai Peddireddy, Benjamin Hurt, Jiangzhuo Chen, Anil Vullikanti, Madhav Marathe, Stefan Hoops, Parantapa Bhattacharya, Dustin Machi, Shi Chen, Rajib Paul, Daniel Janies, Jean-Claude Thill, Marta Galanti, Teresa Yamana, Sen Pei, Jeffrey Shaman, Nicholas G. Reich, Jessica M. Healy, Rachel B. Slayton, Matthew Biggerstaff, Michael A. Johansson, Michael C. Runge, Cécile Viboud

## Abstract

**What is already known about this topic?:** The highly transmissible SARS-CoV-2 Delta variant has begun to cause increases in cases, hospitalizations, and deaths in parts of the United States. With slowed vaccination uptake, this novel variant is expected to increase the risk of pandemic resurgence in the US in July—December 2021.

**What is added by this report?:** Data from nine mechanistic models project substantial resurgences of COVID-19 across the US resulting from the more transmissible Delta variant. These resurgences, which have now been observed in most states, were projected to occur across most of the US, coinciding with school and business reopening. Reaching higher vaccine coverage in July—December 2021 reduces the size and duration of the projected resurgence substantially. The expected impact of the outbreak is largely concentrated in a subset of states with lower vaccination coverage.

**What are the implications for public health practice?:** Renewed efforts to increase vaccination uptake are critical to limiting transmission and disease, particularly in states with lower current vaccination coverage. Reaching higher vaccination goals in the coming months can potentially avert 1.5 million cases and 21,000 deaths and improve the ability to safely resume social contacts, and educational and business activities. Continued or renewed non-pharmaceutical interventions, including masking, can also help limit transmission, particularly as schools and businesses reopen.

## Overview

The rapid development, scale-up, and deployment of COVID-19 vaccines in the United States (US) has been one of the biggest public health successes in the US during this pandemic, with reported cases in June 2021 at the lowest level since mid-March 2020,^1^ despite increased testing capacities. With this success, non-pharmaceutical interventions (NPIs) were lifted, including mask mandates, in almost every jurisdiction across the US. However, the emergence of novel variants with increased transmissibility, particularly the Delta variant, raised concern about the potential timing and magnitude of the resurgence in the coming months, and the ability to mitigate it through increased uptake of vaccination. The COVID-19 Scenario Modeling Hub uses a multiple-model approach to project the state and national trajectories of cases, hospitalizations, and deaths in the US across four scenarios over a six-month time horizon.^2^ As part of the most recent two rounds of projections (Round 6 and 7), increased transmissibility variants were incorporated into projections to assess the potential impact of the Delta variant. In all scenarios with the highest variant transmissibility (60% increase over the Alpha variant, which most closely resembles estimates for the Delta variant), cases were projected to increase in early July 2021 at the national level and peak in mid to late September 2021. Corresponding increases in hospitalizations and deaths were also projected. In scenarios with higher vaccine coverage, the size and duration of this resurgence was notably smaller. This resurgence was projected to be geographically heterogeneous; although most states are projected to experience some degree of rebound, those having higher vaccine coverage are projected to experience less severe increases in incidence relative to prior observed peaks. As of August 2021, reported cases in the US are currently tracking with or above the higher variant transmissibility scenarios. Resurgences were also projected in scenarios with 40% increased transmissibility, although these resurgences are substantially decreased and the peak is delayed, occurring in early October 2021. In contrast, in low increased transmissibility variant scenarios (20% increase over Alpha in Round 6), the epidemic was projected to continue on a downward trajectory in all US jurisdictions.

## Methods

The COVID-19 Scenario Modeling Hub^3^ convened nine modeling teams in an open call to provide six-month (July 3, 2021-January 1, 2022) COVID-19 projections in the US using data available through July 3, 2021. Each team developed a model to project weekly reported cases, hospitalizations, and deaths, both nationally and by jurisdiction (50 states and the District of Columbia), for four different epidemiological scenarios. Models were calibrated against data from the Johns Hopkins Center for Systems Science and Engineering Coronavirus Resource Center and federal databases.^4,5^ The four scenarios included low and high vaccination hesitancy levels, assuming national vaccination coverage saturation at 80% and 70%, respectively, based on hesitancy surveys (Table).^6,7^ Participating teams accounted for vaccination rates by state, age, and risk-groups (e.g., older adults and health care workers). Specified vaccine efficacy levels were constant across the scenarios and were based on protection against clinical disease in randomized clinical trials and effectiveness studies; parameters for effectiveness against infection, transmission, and progression to severe outcomes (e.g., death) were left to be specified by each team.^3^

**TABLE.**
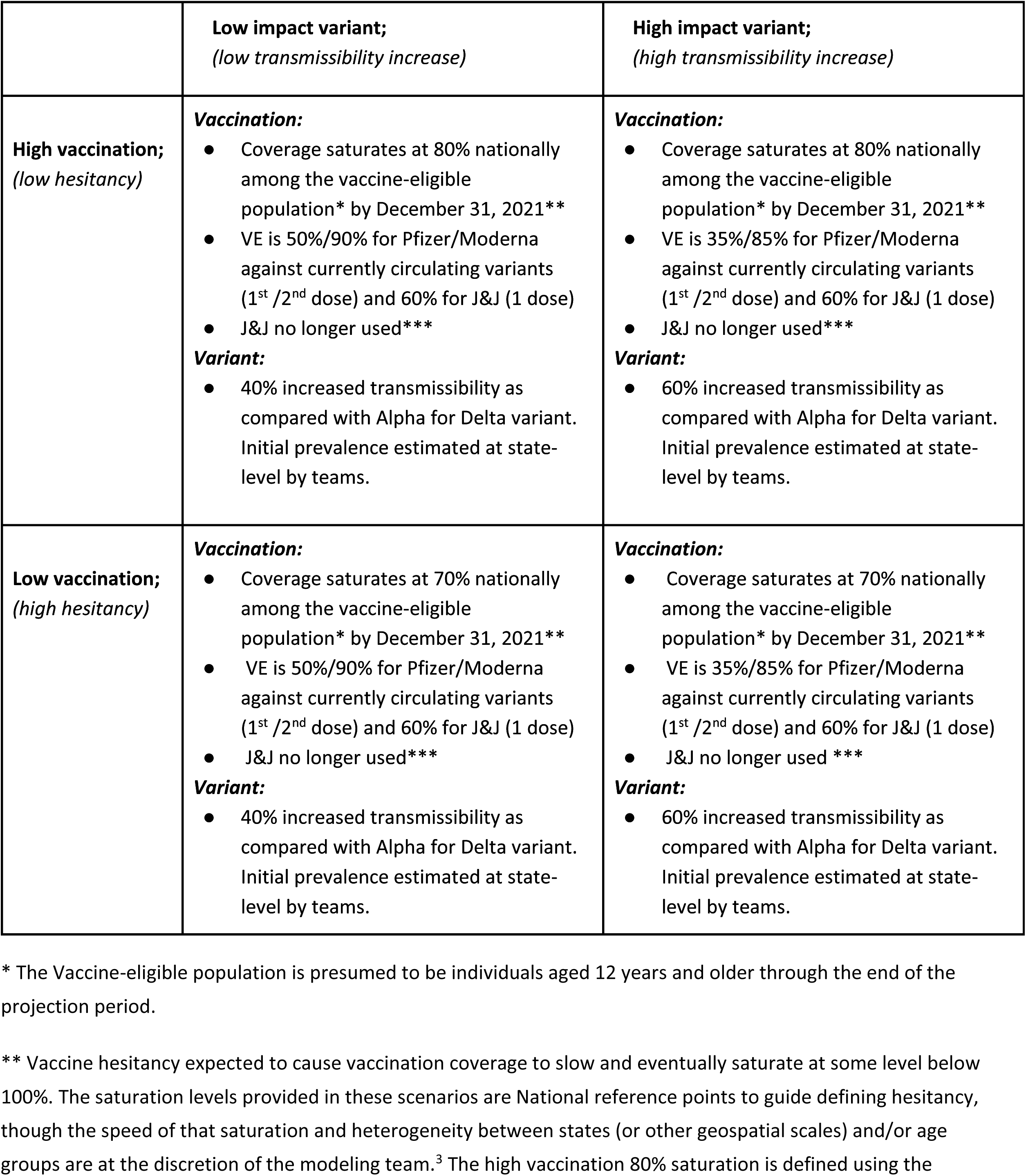
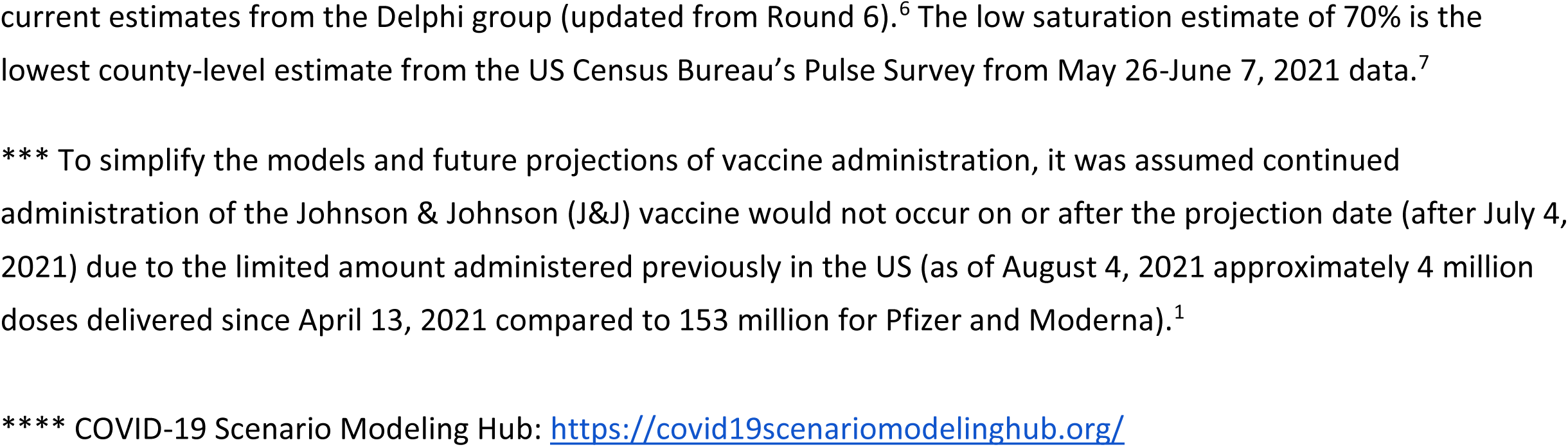
COVID-19 projection scenarios* — United States, July 4, 2021–January 1, 2022. Scenarios defined for projection of COVID-19 cases, hospitalizations, and deaths for the sixth round of projections through the COVID-19 Scenario Modeling Hub****.

|  | **Low impact variant;**<br>*(low transmissibility increase)* | **High impact variant;**<br>*(high transmissibility increase)* |
| --- | --- | --- |
| **High vaccination;**<br>*(low hesitancy)* | **Vaccination:**<br>Coverage saturates at 80% nationally among the vaccine-eligible population* by December 31, 2021**<br>VE is 50%/90% for Pfizer/Moderna against currently circulating variants (1<sup>st</sup> /2<sup>nd</sup> dose) and 60% for J&J (1 dose)<br>J&J no longer used***<br>**Variant:**<br>40% increased transmissibility as compared with Alpha for Delta variant. Initial prevalence estimated at state-level by teams. | **Vaccination:**<br>Coverage saturates at 80% nationally among the vaccine-eligible population* by December 31, 2021**<br>VE is 35%/85% for Pfizer/Moderna against currently circulating variants (1<sup>st</sup> /2<sup>nd</sup> dose) and 60% for J&J (1 dose)<br>J&J no longer used***<br>**Variant:**<br>60% increased transmissibility as compared with Alpha for Delta variant. Initial prevalence estimated at state-level by teams. |
| **Low vaccination;**<br>*(high hesitancy)* | **Vaccination:**<br>Coverage saturates at 70% nationally among the vaccine-eligible population* by December 31, 2021**<br>VE is 50%/90% for Pfizer/Moderna against currently circulating variants (1<sup>st</sup> /2<sup>nd</sup> dose) and 60% for J&J (1 dose)<br>J&J no longer used***<br>**Variant:**<br>40% increased transmissibility as compared with Alpha for Delta variant. Initial prevalence estimated at state-level by teams. | **Vaccination:**<br>Coverage saturates at 70% nationally among the vaccine-eligible population* by December 31, 2021**<br>VE is 35%/85% for Pfizer/Moderna against currently circulating variants (1<sup>st</sup> /2<sup>nd</sup> dose) and 60% for J&J (1 dose)<br>J&J no longer used ***<br>**Variant:**<br>60% increased transmissibility as compared with Alpha for Delta variant. Initial prevalence estimated at state-level by teams. |

\* The Vaccine-eligible population is presumed to be individuals aged 12 years and older through the end of the projection period.
\*\* Vaccine hesitancy expected to cause vaccination coverage to slow and eventually saturate at some level below 100%. The saturation levels provided in these scenarios are National reference points to guide defining hesitancy, though the speed of that saturation and heterogeneity between states (or other geospatial scales) and/or age groups are at the discretion of the modeling team.<sup>3</sup> The high vaccination 80% saturation is defined using the
current estimates from the Delphi group (updated from Round 6).<sup>6</sup> The low saturation estimate of 70% is the lowest county-level estimate from the US Census Bureau's Pulse Survey from May 26-June 7, 2021 data.<sup>7</sup>
\*\*\* To simplify the models and future projections of vaccine administration, it was assumed continued administration of the Johnson & Johnson (J&J) vaccine would not occur on or after the projection date (after July 4, 2021) due to the limited amount administered previously in the US (as of August 4, 2021 approximately 4 million doses delivered since April 13, 2021 compared to 153 million for Pfizer and Moderna).<sup>1</sup>

Scenarios assumed one of two levels of increased transmissibility for the Delta variant: 40% (low) or 60% (high) more transmissible than the Alpha variant. The previous round (Round 6) also included a 20% increase in transmissibility as one scenario option (Supplemental Table 1). Increases in new variant prevalence over time were determined by each modeling team and were estimated at the state level. Individual modeling teams provided probabilistic projections of epidemic trajectory for each week of the projection period. These individual projections were combined into an ensemble for each scenario, outcome, week, and location using an equally-weighted linear opinion pool method across teams that trimmed the highest and lowest model at each point.^8,9^ Individual models differed substantially in structure and design, the details of which can be accessed through the COVID-19 Scenario Modeling Hub website.^10^ Point estimates provided here are the median of the ensemble. For any given pair of scenarios, averted cases and deaths were calculated as the difference (and ratio) between the median point estimates of the ensemble for the two scenarios. To provide a relative measure of resurgence in each state, we compared the intensity of the projected outbreak in the next 6 months to the size of the winter 2020-2021 outbreak -- a period of high hospital burden in many jurisdictions. Specifically, projected resurgences were assessed by taking the ratio of the peak projected median incidence in a given location to the highest incidence experienced during the winter 2020-2021 period (defined as October 1, 2020–February 28, 2021) for the same location. Winter 2020-21 peaks were identified as the 7-day average centered around the day with the highest incident cases from smoothed curves generated through a penalized cubic spline Poisson regression model fit to the incident cases.

## Results

In the two scenarios with high Delta variant transmissibility (60% more transmissible than Alpha), we projected a national wave of cases to continue to grow over the summer and peak in mid-to late September 2021. In the scenario that assumes lower vaccination coverage among eligible individuals (70%) and higher variant transmissibility (the most pessimistic scenario), this resurgence is projected to peak at 414,000 weekly cases (95% projection interval (PI): 103,000–1,525,000) and 5,900 weekly deaths (95% PI: 900-30,000) nationally. Overall, this scenario projects 7,554,000 (95% PI: 2,279,000– 28,399,000) cumulative cases and 96,000 (95% PI: 27,000–476,000) cumulative deaths during July 4, 2021–Jan 1, 2022 (Figure 1).

**FIGURE 1.**
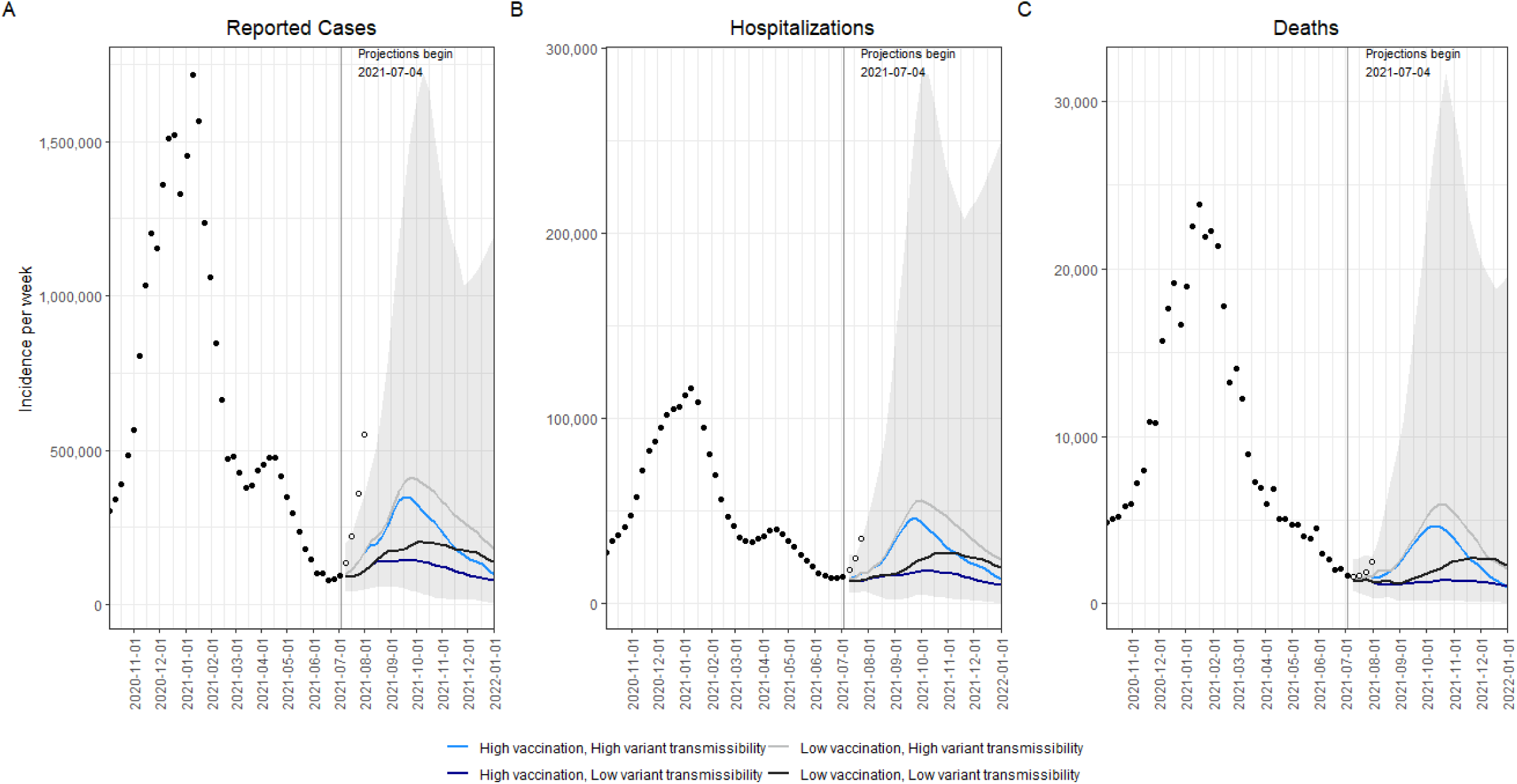
Historical data and weekly ensemble projections of reported numbers of COVID-19 cases (A), hospitalizations (B), and deaths (C) under four scenarios representing different levels of vaccination and Delta variant transmissibility increase — United States, October, 2020–December, 2021. Projections are ensemble estimates of 9 models projecting four 6-month scenarios with 95% prediction intervals (the grey shading encompasses the prediction intervals from all four scenarios). Projections used empirical data from up to July 3, 2021, to calibrate models. The vertical lines indicate the beginning of each projection, only data available prior to that point were used to fit the projections.

With higher variant transmissibility, increasing national vaccination coverage is projected to temper the fall wave slightly and cause it to drop more quickly, but not prevent it. With an increase in national vaccine coverage to 80% by January 1, 2022, the ensemble projected 65,000 (17%) fewer cases and 1,300 (22%) fewer deaths per week at the peak, and 1,525,000 (20%) and 21,000 (22%) fewer cumulative cases and deaths, respectively, during July 4, 2021–January 1, 2022, when compared to the scenario where vaccination saturated at 70% nationally (Figure 1).

The projected national resurgence in COVID-19 cases in the higher transmissible variant scenarios was composed of highly heterogeneous state-level resurgences. The ten states with the largest projected increases in incidence relative to their winter 2020-21 peak are, in descending order, Louisiana, Hawaii, Nevada, Arkansas, Missouri, Florida, Utah, Georgia, Alabama, and Mississippi. The ensemble estimates project these states to experience median peak levels of weekly incident cases that are 19–71% (95% PI: 1-534%) of their (smoothed) winter peak, although this has already been exceeded in some states (e.g., Florida, Louisiana). The ten states with the lowest projected resurgences are, in ascending order, Massachusetts, Minnesota, Rhode Island, Pennsylvania, North Dakota, Wisconsin, Vermont, Maine, Tennessee, and New York. The ten states with the least resurgence had a median first-dose vaccine coverage of 71% among the eligible population (ages 12+) on July 3, 2021, compared to 52% in the ten states with the highest projected resurgence. We find a high negative correlation (Pearson’s *r* = -0.66, Figure 2) between projected *cumulative deaths* per population and vaccination coverage on July 3, 2021.^11^ In all states, even those with low overall vaccination coverage, at least 76% of people 65+ had received at least one dose of the vaccine, which was expected to have a major effect in limiting mortality from Delta.^1^ The impact of vaccination is already being observed: in the ten states with the largest projected resurgence there has been a 9% reduction in the observed case fatality ratio (CFR) comparing August-December 2020 and January-July 2021; in the ten states with the least projected resurgence a 21% reduction in CFR has been observed. During the projection period, we project reductions of 15% and 14%, as compared to August-December 2020.

**FIGURE 2.**
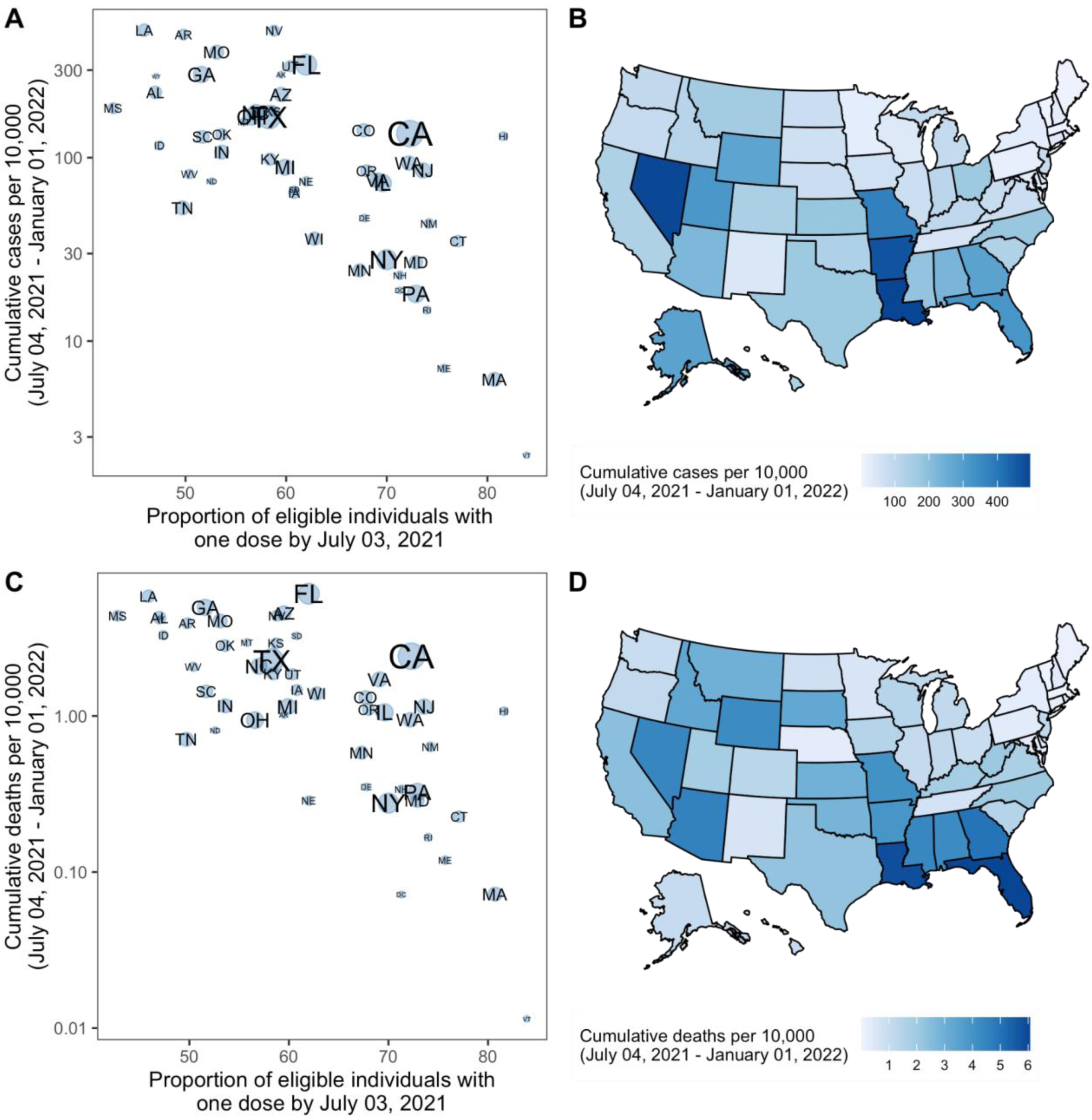
Projected cumulative cases and mortality in the most pessimistic scenario (low vaccination, high variant transmissibility) and current vaccination coverage by state — United States, July 4 2021– January 1, 2022. (A) Correlation between cumulative projected cases per 10,000 population during the 6-month period and proportion of the eligible population vaccinated with at least one COVID-19 vaccine dose by July 3, 2021, by state. Circle sizes represent population size. (B) Cumulative projected cases per 10,000 population during the 6-month period, by state. (C) Correlation between cumulative projected deaths per 10,000 population during the 6-month period and proportion of the eligible population vaccinated with at least one COVID-19 vaccine dose by July 3, 2021, by state. Circle sizes represent population size. (D) Cumulative projected deaths per 10,000 population during the 6-month period, by state.

Lower transmissibility variant scenarios project significantly reduced resurgence, projecting cumulative national cases of only 9 and 13% compared to the winter 2020-21 peak. In the previous projection round (Round 6), resurgence was expected to produce only 8% of the cases reported during the winter 2020-21 peak nationally in scenarios assuming only a 20% transmissibility increase from a novel variant.^3^

As of July 31, 2021, reported cases in the US exceeded those projected in all scenarios, including those assuming 60% increased transmissibility of the Delta variant (Figure 1). We compared weekly incident and cumulative cases during the first four weeks after the projection date (July 4-31, 2021). The total median projected number of cases underestimates the observed cases overall during this 4-week period (1,256,000 observed vs 218,000 projected); however, we find a strong correlation between ranking of observed and projected total cases per 100,000 during the first four weeks of the projection period, at the state level (Spearman’s *ρ*= 0.87, Figure 3). Seven of the 10 states with greatest projected incidence rank in the ten worst observed incidence states. These rapid increases in observed cases may not represent the final resurgent sizes, but currently reflect the expected severity ranking among projections well.

**FIGURE 3.**
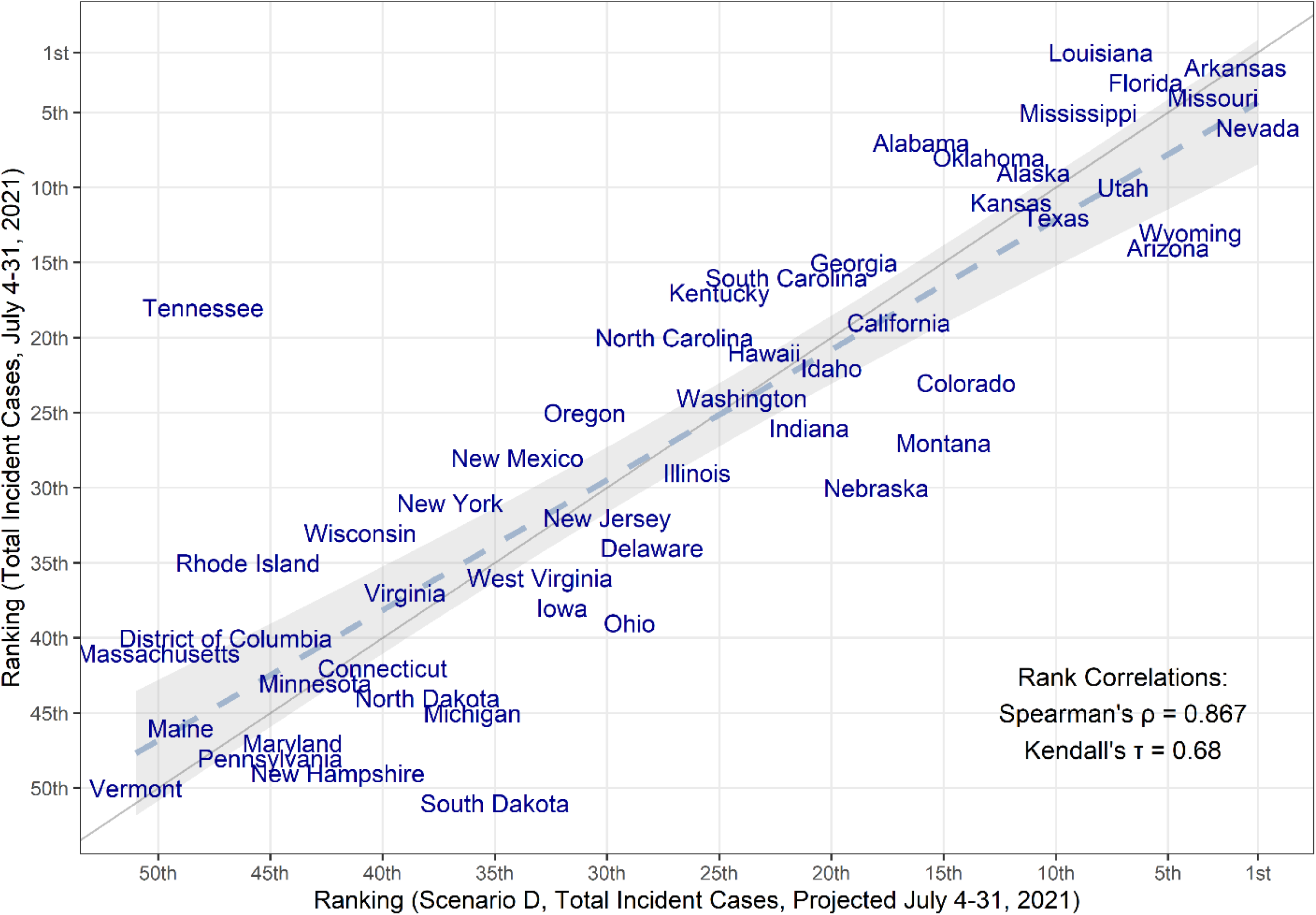
Comparison of the median projected and observed state-level total COVID-19 case incidences occurring during July 4-31, 2021, United States. Comparison is based on ranking of incidence per capita in 50 states + DC (Spearman’s rank correlation=0.867). The grey solid line represents perfect agreement between ranks (y=x), while the dashed line and grey shaded area represent a regression line fitted to the data, with confidence intervals.

## Discussion

The introduction and rapid spread of the SARS-CoV-2 Delta variant in the US, to which 81% of recent infections were attributed as of the two weeks ending July 31, 2021,^1^ has prompted concerns about the scale of the COVID-19 resurgence in the US in the summer and fall of 2021, especially in the midst of decreased NPIs and slowing vaccination rates. Projections combining insights from multiple models suggest sizable resurgences of COVID-19 across the US, assuming growth of a variant that is 60% more transmissible than the Alpha variant (an assumption aligned with most estimates of the relative transmissibility of these variants).^12^ In scenarios with higher vaccination coverage, the magnitude of the resurgence in cases and deaths was substantially lower than in the lower coverage scenarios. Only 13 states and Washington, D.C. accomplished President Biden’s goal for vaccination coverage among eligible populations at or above 70% by July 1, 2021. Efforts to increase vaccination rates will save lives before and during future resurgences and are critical.

The rapid case growth observed in July 2021 in multiple US states has been surprising, tracking with or above the projections from our worst-case scenario. Scenarios were designed at the end of June 2021 based on information available at the time about the transmissibility of the Delta variant, vaccine effectiveness against Delta, and vaccine coverage; projections were generated on or before July 4, 2021. Our data should not be understood as forecasts, but as projections conditional on the scenarios; several reasons could explain why case growth has been faster than expected, particularly if scenarios omitted key epidemiological or behavioral aspects affecting disease dynamics. Possible mechanisms include incomplete assumptions about the transmissibility and severity of Delta (including changes in serial interval or severity of infection relative to other variants), the effectiveness of the vaccines against infection and transmission, changes in testing practices, and the interaction of these factors with NPIs and behavior change.^13–15^ Use of NPIs has been substantially reduced across the US, with a lapse in mask mandates in most states. Although modeled use of NPIs was left to the discretion of individual modeling teams and did not vary between scenarios, results of prior rounds underscore the effectiveness of NPIs, in combination with increasing vaccination, to moderate the spread of a highly transmissible variant.^2^ The extent to which NPIs may be necessary will vary across states, as those states with high levels of vaccination coverage or natural immunity are at lower risk for an increase in cases. Widespread implementation of a recent change in CDC recommendations regarding mask mandates may help reduce transmission.^16^

The impact of the resurgence on severe disease and mortality is expected to vary substantially across states; states with younger populations and higher vaccination coverage among older and high-risk populations are expected to experience relatively lower burden of severe disease, even with resurgences in cases. Several states (e.g., South Dakota, North Dakota) with low vaccination coverage were not projected to experience major resurgence, likely because of high naturally acquired immunity. In addition, as the projected resurgence may continue throughout the summer and into the beginning of the school year, efforts to promote vaccination among eligible school-aged children and college students and to maintain key prevention strategies in schools (e.g., mask-wearing among the unvaccinated, physical distancing, screening programs) can help reduce risks with a safe return to in-person instruction.^16^ A recent surge in new vaccinations, particularly among young age groups and in jurisdictions most severely impacted by the Delta variant, is a positive step in this direction.^1^

The findings in this report are subject to several limitations. First, considerable uncertainty is inherent to long-term projections. This has been repeatedly illustrated throughout the COVID-19 pandemic, with rapid changes in behavior, deployment of vaccines and boosters, and the emergence of novel variants, each of which has the capacity to drastically shift the epidemic trajectories. Uncertainty may arise from three main sources: specification of the scenarios (e.g., uncertainty in transmissibility); errors in the structure or assumptions of individual models given a specific scenario (e.g., variations in assumptions about vaccination uptake); and inaccurate calibration based on incomplete or biased data (e.g., reporting backlogs). None of the 4 scenarios considered here are likely to precisely reflect the future reality over a 6-month period.^2^ Further, for a given scenario, there is notable variation among individual model projections with regards to both the timing and the magnitude of the resurgence. Variation likely reflects differences in model structure, projected vaccine coverage, projected variant growth, and importance of seasonal effects. In addition, these scenarios do not specify considerations of Delta infecting previously immune individuals, the waning of existing immunity, increases in NPIs, or vaccination among children aged <12 years, all of which may be important drivers of dynamics in the coming months. In the same vein, model estimates are dependent on assumptions about vaccine hesitancy, which are informed in part by large-scale surveys of vaccine sentiments.^6,7^ These surveys may underestimate vaccine hesitancy, as current coverage estimates among survey respondents are substantially higher than measured among the overall US population. Additionally, there are limitations to individual component models, though these concerns are tempered by analyzing ensembles of the nine different models.

## Conclusions

The emergence and introduction of more transmissible SARS-CoV-2 variants like the Delta variant are projected to lead to a substantial resurgence of COVID-19 in the US and are already causing increasing cases in every state across the country. The high variant transmissibility scenarios, which appear to more accurately represent the characteristics of the Delta variant, both in transmissibility and in current case trajectories, projected a significant national resurgence with substantial variation in magnitude across states. Resurgences are expected to be more pronounced in low-vaccination jurisdictions. The projections indicate that even with substantial vaccination coverage, the increased transmissibility of new variants like Delta can continue to challenge our ability to control this pandemic. Renewed efforts to increase vaccination coverage are critical to limiting transmission and disease, particularly in states with low natural immunity and lower current vaccination, in addition to re-instituting control measures like indoor masking when needed. Multi-model ensemble efforts such as the COVID-19 Scenario Modeling Hub are particularly well-suited to provide disease projections to inform the pandemic response under changing epidemiological and behavioral situations.

## Data Availability

All projection results are publicly available. Other data referred to in the manuscript are from publicly available sources cited in the manuscript.

https://covid19scenariomodelinghub.org/

https://github.com/midas-network/covid19-scenario-modeling-hub

## Data Availability

https://covid19scenariomodelinghub.org/

https://github.com/midas-network/covid19-scenario-modeling-hub

## Disclaimer

The findings and conclusions in this report are those of the authors and do not necessarily represent the views of the Centers for Disease Control and Prevention or the National Institutes of Health. Any use of trade, firm, or product names is for descriptive purposes only and does not imply endorsement by the US Government.

## Acknowledgements

IHME (Bobby Reiner) for helpful discussions.

## Notes

### Competing Interest Statement

JL has served as an expert witness on cases where the likely length of the pandemic was of issue. MCR reports stock ownership in Becton Dickinson & Co., which manufactures medical equipment used in COVID-19 testing, vaccination, and treatment. JS and Columbia University disclose partial ownership of SK Analytics. JS discloses consulting for BNI.
there are no other competing interests to declare.

### Funding Statement

Katriona Shea reports receipt of two National Science Foundation (NSF) COVID-19 RAPID awards, and a Huck Institutes of the Life Sciences Coronavirus Research Seed Grant. Rebecca Borchering reports funding from an NSF COVID-19 RAPID award. Katharine Tallaksen, Kaitlin Rainwater-Lovett, Laura Asher, Luke C. Mullany, Molly E. Gallagher, Matt Kinsey, Richard F. Obrecht, and Lauren Shin report funding from the U.S. Department of Health and Human Services (HHS), Office of the Assistant Secretary for Preparedness and Response to the Johns Hopkins Applied Physics Laboratory. Matteo Chinazzi reports grants from the National Institutes of Health (NIH), the Council of State and Territorial Epidemiologists (CSTE), and Metabiota to Northeastern University. Ana Pastore y Piontti reports funding from Metabiota, Inc. to Northeastern University and royalties from Springer Publishing. Joseph Lemaitre reports funding from the Swiss National Science Foundation, State of California, HHS, and the Department of Homeland Security (DHS). Kyra H. Grantz reports support from the California Department of Public Health, Johns Hopkins Bloomberg School of Public Health, NIH, and travel support from the World Health Organization (WHO). Elizabeth Lee and Claire Smith report support from the California Department of Public Health, Johns Hopkins Bloomberg School of Public Health, Johns Hopkins Health System, HHS, and DHS, and computing resources from Amazon Web Services, Johns Hopkins University Modeling and Policy Hub, and the Office of the Dean at the Johns Hopkins Bloomberg School of Public Health. Justin Lessler reports support from DHHS, DHS, California Institute of Technology, NIH, honorarium from the American Association for Cancer Research, personal fees for expert testimony from Paul, Weiss, Rifkind, Wharton & Garrison, LLP. Lindsay Keegan reports support from the State of California, and NIH, a University of Utah Immunology, Inflammation, and Infectious Disease Seed Grant, and a scholarship from the University of Washington Summer Institute in Statistics and Modeling of Infectious Diseases. Lucie Contamin, John Levander, Jessica Salerno, and Harry Hochheiser report a National Institute of General Medical Sciences grant (U24GM13201). Ajitesh Srivastava reports a grant from the National Science Foundation. Alessandro Vespignani reports grants from NIH, NSF, WHO, CSTE, Metabiota Inc., Templeton Foundation, Scientific Interchange Foundation, Bill & Melinda Gates Foundation; royalties from Cambridge University Press, World Scientific, Springer Publishing, and Il Saggiatore; consulting fees from Human Technopole Foundation, Institute for Scientific Interchange Foundation, honorarium for lecture module at University of Washington; Scientific Advisory Board member of the Institute for Scientific Interchange Foundation, Italy, Supervisory Board member of the Human Technopole Foundation, Italy; and gifts to Northeastern University from the McGovern Foundation, the Chleck Foundation, the Sternberg Family, J. Pallotta, and Google Cloud research credits for COVID-19 from Google. Akhil Sai Peddireddy, Pyrros A. Telionis, Anil Vullikanti, Jiangzhuo Chen, Benjamin Hurt, Brian D. Klahn, Bryan Lewis, James Schlitt, Joseph Outten, Lijing Wang, Madhav Marathe, Patrick Corbett, Przemyslaw Porebski, and Srinivasan Venkatramanan report institutional support from the National Science Foundation, Expeditions, NIH, the U.S. Department of Defense, Virginia Department of Health, Virginia Department of Emergency Management, University of Virginia (internal seed grants), and Accuweather. No other potential conflicts of interest were disclosed.

### Author Declarations

This analysis was conducted using publicly available, aggregated data on cases and deaths, along with published parameter estimates. No human subjects research was involved, and no IRB approval was required.

### Summary of Updates

Added funding and COI statements. Removed one author.

